# Polygenic score for physical activity provides odds for multiple common diseases

**DOI:** 10.1101/2021.02.12.21251632

**Authors:** Elina Sillanpää, Teemu Palviainen, Finn gen, Samuli Ripatti, Urho M. Kujala, Jaakko Kaprio

**Author notes:** Corresponding author: Elina Sillanpää, PhD, Gerontology Research Center, Faculty of Sport and Health Sciences, P.O. Box 35 (VIV), FIN-40014 University of Jyväskylä, Finland.

## Abstract

**Purpose:** It has been suggested that genetic pleiotropy, in which the same genes affect two or more traits, may partially explain the frequently observed associations between high physical activity (PA) and later reduced morbidity or mortality. However, the evidence about pleiotropy from human studies is limited. This study investigated associations between PA polygenic risk scores (PRSs) and cardiometabolic diseases among the Finnish population.

**Methods:** PRSs for device-measured overall PA were adapted to a FinnGen study cohort of 218,792 individuals with genome-wide genotyping and extensive digital longitudinal health register data. Associations between PA PRS and body mass index (BMI), diseases, and mortality were analysed with linear and logistic regression models. The number of different disease endpoints varied between 894 and 111,108 in FinnGen cohort.

**Results:** A high PA PRS predicted a lower BMI (β −0.025 kg/m^2^ per one standard deviation (SD) change in PA PRS, SE 0.013, p=1.87×10^−80^). The PA PRS also predicted a lower risk for diseases that typically develop later in life or not at all among highly active individuals. A lower disease risk was systematically observed for cardiovascular diseases [odds ratio, OR per 1 SD change in PA PRS 0.95, p=9.5*10^−19^) and, for example, hypertension [OR 0.93, p=2.7*10^−44^), type 2 diabetes (OR 0.91, p=4.1*10^−42^), and coronary heart disease (OR 0.95 p=1.2*10^−9^). Participants with high PA PRS had also lower mortality risk (OR 0.97, p=0.0003). We did not observe statistically significant associations with hypothetical control conditions, such as osteoarthritis and osteoporosis.

**Conclusions:** Genetically less active persons are at a higher risk of developing cardiometabolic diseases, which may partly explain the previously observed associations between low PA and higher disease and mortality risk. The same inherited physical fitness and metabolism related mechanisms may be associated both with PA levels and with cardiometabolic disease risk.

## INTRODUCTION

Non-communicable diseases, such as cardiovascular disease, type 2 diabetes, and neoplasms, cause a large burden to society, with an estimated one billion cases existing worldwide. Therefore, early detection, prevention, and intervention regarding these diseases are fundamental goals in advancing human health and quality of life [1]. To date, it is known that individual disease risk is a complex interplay of genetic susceptibility and multiple social, environmental, and policy factors.

Genetic risk estimate is the earliest measurable contributor to overall disease risk during person”s life span. Genetic contributions to complex traits and diseases, such as physical activity (PA) and cardiometabolic diseases (CMDs), are polygenic (i.e., accounted for by a large number of causal variants with very small effects). Polygenic risk scores (PRSs) summarize genome-wide genotype data into single variables that produce individual-level risk scores regarding genetic liability. PRSs already have been produced for several CMD traits [2-4]. These CMD PRSs have confirmed the existence of genetic influence on common disease risks previously reported in twin and family studies [2]. CMD PRSs have improved the clinical risk prediction of CMDs and predicted disease onset—especially among high-risk individuals [5]. In contrast to CMDs, PRSs for lifestyle factors are less frequently used. Multiple twin studies suggest that human behaviour is moderately genetically regulated [6], and PRSs can be calculated to any heritable trait. In year 2020 we published two PRSs for PA and showed their significant out-of-sample predictive values in two independent cohorts with different PA phenotypes [7].

The existence of polygenic influences on both PA and CMD suggests that gene–environment interplay and reverse causality may play a role in associations between PA, CMD, and mortality [8-11]. PA has been highlighted as a cost-effective strategy for the prevention of CMDs, as observational epidemiological studies consistently show that high PA levels strongly predict lower disease risk and all-cause and cause-specific mortality. Clinical trials and field interventional studies have not provided strong evidence for or against a causal role of PA in mortality due to lack of long-term follow-up. PA has generally beneficial—but relatively modest—effects on biological risk factors for disease, such as improvements in blood lipid levels, blood pressure and glucose metabolism.

The latest animal and human findings challenge the assumption regarding the causal association between higher PA and reduced mortality risk later in life [12, 13]. When genetic factors are fully controlled, twin studies suggest that PA does not reduce mortality risk [12]. It has been suggested that genetic pleiotropy, where the same genes affects two or more characters, may partially explain the frequently observed associations between high PA and reduced mortality risk later in life [8, 12], but evidence from human studies is limited.

Genetic confounding occurs when a genetic variant or set of variants causally affect both the risk factors and outcomes (e.g., variants associated with CMD risk factors, such as PA, also directly affect CMD). This causes challenges in observational epidemiology, as adjusting for genetic confounders is typically insufficient in analysis [14]. Another potential source of bias is gene–environment interaction. Although the impact of genetic inheritance on PA is generally poorly understood, it is assumed that individuals with favourable genotypes tend to participate in physical activities. Studies suggest that these individuals may have inherited better cardiorespiratory fitness [15], derive greater pleasure from physical activities, and that their personality and other behavioural characteristics makes it easier for them to adopt and follow a physically active lifestyle [16]. This gene–environment interaction hinders causal reasoning because the environment or lifestyle experienced by an individual is partly influenced by their genotypes. The PRS construct offers tools to answer confounding and gene-environment challenges in observational research. Individual PRSs can be used to explore potential genetic overlap in two or more traits (such as PA and CMDs) as well as to predict other traits in a regression model across a study sample.

This study investigated associations between PA PRS, CMD and mortality among the Finnish population study cohort FinnGen of 218,792 individuals.

## RESULTS

A brief description of the study design is presented in Figure 1. We observed that a high PA PRS, a genetic inheritance that supports higher volumes for physical activity, systematically predicted a lower risk for diseases that typically develop later in life or not at all among highly active individuals.

**Figure 1.**
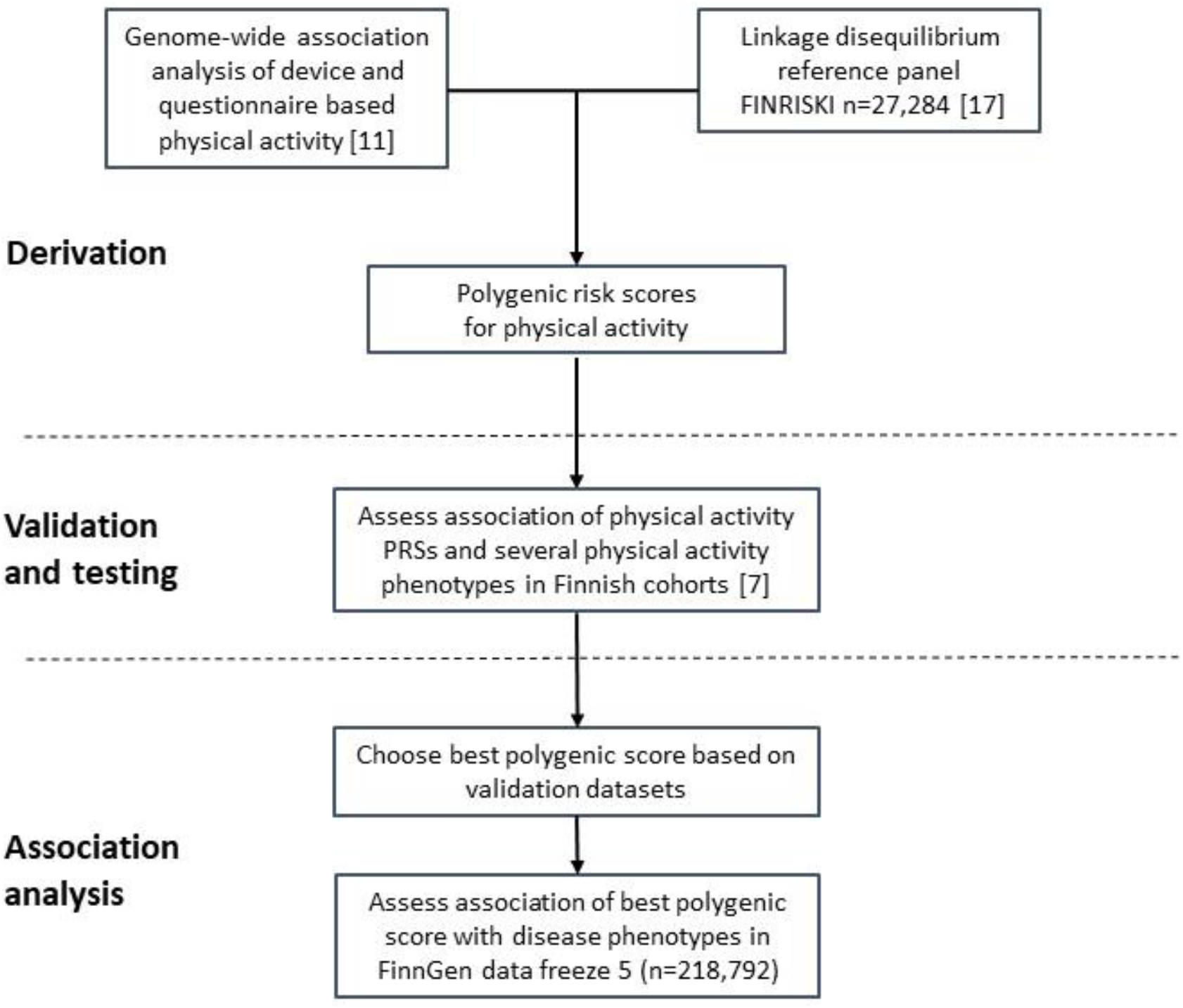
Study design and workflow. Polygenic risk scores for physical activity were derived from recent GWA studies ([11] and http://www.nealelab.com) and a linkage disequilibrium reference panel of 27,284 unrelated Finnish individuals [17]. Two polygenic scores for physical activity were derived and their out-of-sample predictive values were tested using two independent Finnish cohorts and several physical activity phenotypes [7]. The polygenic score for device-based measures of overall physical activity volume was selected for association analysis, which was conducted in a FinnGen cohort of 218,792 Finnish participants. The clinical endpoints utilized in the analysis were derived from Finnish nation-wide digital health registers.

### Polygenic risk, BMI, and obesity

High PA PRS predicted lower BMI (β −0.025 kg/m2 per standard deviation (SD) of PA PRS, SE 0.013, p=1.87×10^−80^, n=160,334) and body weight (β −0.876 kg per SD of PA PRS, SE 0.041, p=3.69×10^−102^, n=164,964). Higher PA PRS also predicted a decreased risk for obesity (Table 1).

**Table 1.** Association analysis between polygenic score for physical activity and metabolic endpoints as well as selected control conditions.

| Phenotype | N cases/controls | OR | P | BETA | SE |
| --- | --- | --- | --- | --- | --- |
| Obesity | 8,908/209,827 | 0.90 | $5.8 \times 10^{-20}$ | -0.1011 | 0.0108 |
| Type 2 diabetes | 29,193/182,573 | 0.91 | $4.1 \times 10^{-42}$ | -0.0906 | 0.0066 |
| Type 2 diabetes without complications | 14,622/183,185 | 0.91 | $1.0 \times 10^{-27}$ | -0.0978 | 0.0088 |
| Type 2 diabetes with complications | 24,133/183,185 | 0.91 | $1.9 \times 10^{-42}$ | -0.0984 | 0.0071 |
| Type 2 diabetes with peripheral circulatory complications | 1,049/183,185 | 0.91 | 0.0027 | -0.0987 | 0.0310 |
| Type 2 diabetes medication (all types) | 32,897/185,820 | 0.92 | $5.3 \times 10^{-40}$ | -0.0822 | 0.0061 |
| Type 2 diabetes medication (other than insulin) | 28,493/185,895 | 0.91 | $2.7 \times 10^{-44}$ | -0.0938 | 0.0066 |
| Diabetes, insulin treatment (Kela reimbursement) | 29,071/189,721 | 0.92 | $2.8 \times 10^{-36}$ | -0.0820 | 0.0064 |
| Hypothyroidism (congenital or acquired) | 26,342/59,827 | 0.92 | $1.8 \times 10^{-29}$ | -0.0803 | 0.0085 |
| Hypothyroidism, strict autoimmune | 22,997/175,475 | 0.96 | $1.9 \times 10^{-8}$ | -0.0416 | 0.0072 |
| Disorders of lipoprotein metabolism/other lipidaemias | 14,010/197,259 | 0.96 | $8.3 \times 10^{-6}$ | -0.0413 | 0.0090 |
| Hyperlipidaemia, other/unspecified | 4,535/197,259 | 0.94 | 0.0004 | -0.0566 | 0.0152 |
| Nonalcoholic fatty liver disease | 894/217,898 | 0.89 | 0.0014 | -0.1128 | 0.0335 |
| Cholelithiasis | 19,023/195,144 | 0.93 | $3.7 \times 10^{-18}$ | -0.0689 | 0.0078 |
| Statin medication | 68,782/150,010 | 0.95 | $3.8 \times 10^{-25}$ | -0.0559 | 0.0053 |
| Sleep apnoea | 16,761/201,194 | 0.93 | $3.7 \times 10^{-20}$ | -0.0762 | 0.0081 |
| Osteopathies and chondropathies | 9,217/209,575 | 1.00 | 0.69 | -0.0050 | 0.0107 |
| Osteoporosis | 3,203/209,575 | 0.99 | 0.62 | -0.0104 | 0.0180 |
| Arthrosis | 37,233/147,221 | 1.00 | 0.78 | 0.0020 | 0.0062 |
| Gonarthrosis | 22,796/147,221 | 0.99 | 0.12 | -0.0128 | 0.0075 |
| Smoking | 29,961/66,872 | 0.95 | $2.8 \times 10^{-9}$ | -0.0507 | 0.0071 |

Logistic regression analysis, OR, odds ratio; SE, standard error. Model adjusted for age, sex, and 10 genetic principal components of population stratification. Kela, the Social insurance Institute of Finland.

### Polygenic risk, type 2 diabetes, and other metabolic diseases

Genetically active participants were at a significantly lower risk for developing type 2 diabetes (OR per SD of PA PRS 0.91, p=4.1*10^−42^). Similar ORs were observed in both uncomplicated type 2 diabetes cases and cases where peripheral complications were present (Table 1).

Higher PA PRS values were also associated with lower odds of diabetes medication and insulin treatment endpoints (based on Kela reimbursement). Genetically active participants also exhibited lower risk for several diseases related to fat metabolism, such as disorders of lipoprotein metabolism, lipidemias, nonalcoholic fatty liver disease, and use of statin medication (ORs from 0.89 to 0.96). Lower odds for hyperthyroidism were also observed among the genetically active participants OR 0.96, p=1.8*10^−29^). However, associations between PA PRS and bone metabolism phenotypes (arthrosis, osteoporosis etc.) were not found. High PA PRS was associated with lower odds for smoking.

### Polygenic risk and cardiovascular diseases

High PA PRS systematically associated with smaller cardiovascular disease (CVD) risk (Table 2).

**Table 2.** Association analysis between polygenic score for physical activity and cardiovascular disease (CVD) endpoints.

| Phenotype | Cases/controls | OR | p-value | beta | SE |
| --- | --- | --- | --- | --- | --- |
| CVD, all | 111,108/107,684 | 0.96 | $9.5 \times 10^{-19}$ | -0.0445 | 0.0049 |
| Coronary atherosclerosis | 23,363/195,429 | 0.95 | $4.4 \times 10^{-11}$ | -0.0514 | 0.0076 |
| Ischemic heart diseases | 30,952/187,840 | 0.96 | $1.1 \times 10^{-10}$ | -0.0447 | 0.0068 |
| Angina pectoris | 18,168/200,624 | 0.95 | $2.5 \times 10^{-10}$ | -0.0540 | 0.0083 |
| Myocardial infarction | 11,622/207,170 | 0.96 | $1.3 \times 10^{-5}$ | -0.0450 | 0.0100 |
| Major CVD event | 21,012/197,780 | 0.95 | $1.2 \times 10^{-9}$ | -0.0491 | 0.0079 |
| Hard CVD | 29,350/189,442 | 0.96 | $4.9 \times 10^{-10}$ | -0.0441 | 0.0069 |
| Coronary revascularization | 12,271/206,521 | 0.93 | $4.7 \times 10^{-13}$ | -0.0685 | 0.0099 |
| All-cause heart failure | 23,397/194,811 | 0.94 | $2.7 \times 10^{-14}$ | -0.0576 | 0.0074 |
| Stroke | 18,661/162,201 | 0.95 | $6.4 \times 10^{-8}$ | -0.0466 | 0.0084 |
| Hypertension | 55,955/162,837 | 0.93 | $2.7 \times 10^{-44}$ | -0.0777 | 0.0055 |
| Antihypertensive medication | 107,287/111,505 | 0.94 | $2.4 \times 10^{-34}$ | -0.0616 | 0.0050 |

Logistic regression analysis, OR, odds ratio; SE, standard error; CVD, cardiovascular disease. Model adjusted for age, sex, and 10 genetic principal components of population stratification.

The genetically active participants had fewer overall CVDs (OR 0.96, p=9.5*10^−19^). They also had a lower risk of ischemic heart disease (OR 0.96 p=1.1*10^−10^), stroke (OR 0.95, p=6.4*10^−8^), and hypertension (OR 0.93, p=2.7*10^−44^), and they also used fewer antihypertensive medications (OR 0.94, p=2.4*10^−34^).

### Polygenic risk, mortality, and dementia

In FinnGen cohort, which included 15,152 deaths, a 1 SD increase in the PA PRS was found to be associated with lower odds for all-cause mortality (OR 0.97, p=0.0003). The risk of Alzheimer’s disease was, however, increased (OR 1.05, p=0.0112), although the number of cases in the FinnGen data was rather modest (n=3,899). The PA PRS was not associated with vascular dementia (Table 3).

**Table 3.** Association analysis between polygenic score for physical activity, dementia endpoints, and death.

| Phenotype | Cases/controls | OR | p-value | beta | SE |
| --- | --- | --- | --- | --- | --- |
| Dementia, all | 7,284/209,487 | 1.01 | 0.4743 | 0.0108 | 0.0128 |
| Alzheimer disease | 3,899/214,893 | 1.05 | 0.0112 | 0.0462 | 0.0170 |
| Alzheimer's disease, atypical or mixed | 800/214,893 | 1.09 | 0.0262 | 0.0863 | 0.0361 |
| Alzheimer's disease, early onset | 587/111,471 | 1.08 | 0.1199 | 0.0724 | 0.0419 |
| Alzheimer's disease, late onset | 2,670/111,471 | 1.02 | 0.3794 | 0.0218 | 0.0216 |
| Vascular dementia | 859/211,300 | 0.99 | 0.7712 | -0.0120 | 0.0346 |
| Any death | 15,152/203,640 | 0.97 | 0.0003 | -0.0335 | 0.0088 |

Logistic regression analysis, OR, odds ratio; SE, standard error. Model adjusted for age, sex, and 10 genetic principal components of population stratification.

**Table 4.**
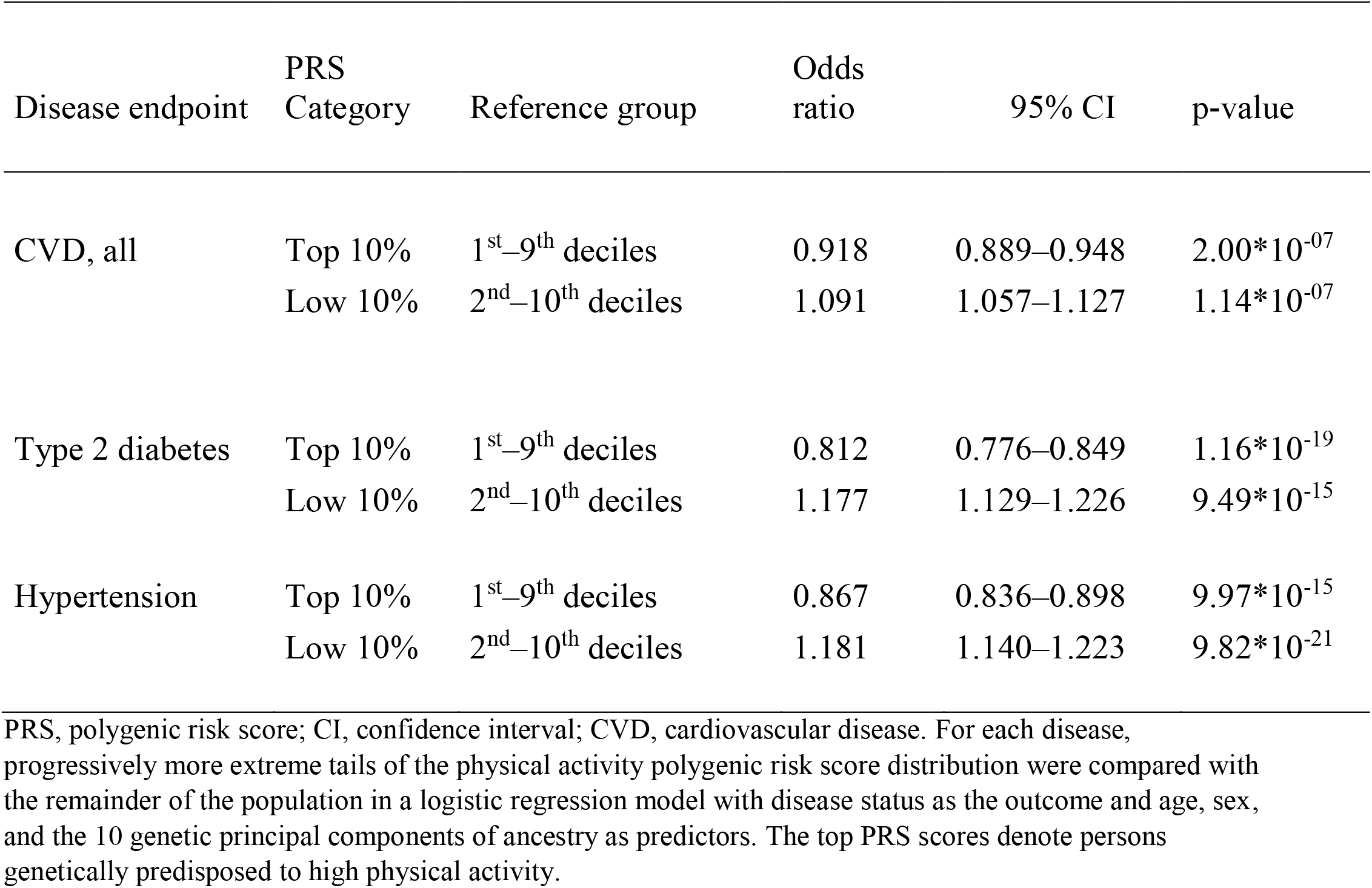
Associations of top and low physical activity polygenic risk scores defined as the top and low deciles of the score distribution with cardiovascular diseases, type 2 diabetes, and hypertension within the FinnGen cohort (n=218,792).

| Disease endpoint | PRS Category | Reference group | Odds ratio | 95% CI | p-value |
| --- | --- | --- | --- | --- | --- |
| CVD, all | Top 10% | 1<sup>st</sup>–9<sup>th</sup> deciles | 0.918 | 0.889–0.948 | $2.00 \times 10^{-07}$ |
| | Low 10% | 2<sup>nd</sup>–10<sup>th</sup> deciles | 1.091 | 1.057–1.127 | $1.14 \times 10^{-07}$ |
| Type 2 diabetes | Top 10% | 1<sup>st</sup>–9<sup>th</sup> deciles | 0.812 | 0.776–0.849 | $1.16 \times 10^{-19}$ |
| | Low 10% | 2<sup>nd</sup>–10<sup>th</sup> deciles | 1.177 | 1.129–1.226 | $9.49 \times 10^{-15}$ |
| Hypertension | Top 10% | 1<sup>st</sup>–9<sup>th</sup> deciles | 0.867 | 0.836–0.898 | $9.97 \times 10^{-15}$ |
| | Low 10% | 2<sup>nd</sup>–10<sup>th</sup> deciles | 1.181 | 1.140–1.223 | $9.82 \times 10^{-21}$ |

PRS, polygenic risk score; CI, confidence interval; CVD, cardiovascular disease. For each disease, progressively more extreme tails of the physical activity polygenic risk score distribution were compared with the remainder of the population in a logistic regression model with disease status as the outcome and age, sex, and the 10 genetic principal components of ancestry as predictors. The top PRS scores denote persons genetically predisposed to high physical activity.

### Analysis of polygenic risk extremities

We further analysed three major CMD phenotypes [cardiovascular diseases (CVD), type 2 diabetes, and hypertension] that were the most strongly associated with high PA PRSs. The median PA PRS percentile score was 50 for participants with CVD and 51 for participants without CVD (Figure 2). The corresponding median value was 48 for the cases and 51 for the controls for type 2 diabetes and 49 and 51 for the hypertensive cases and controls, respectively. For each of the three diseases, we observed a linearly declining trend in disease cases corresponding to increasing PA PRS deciles.

**Figure 2.**
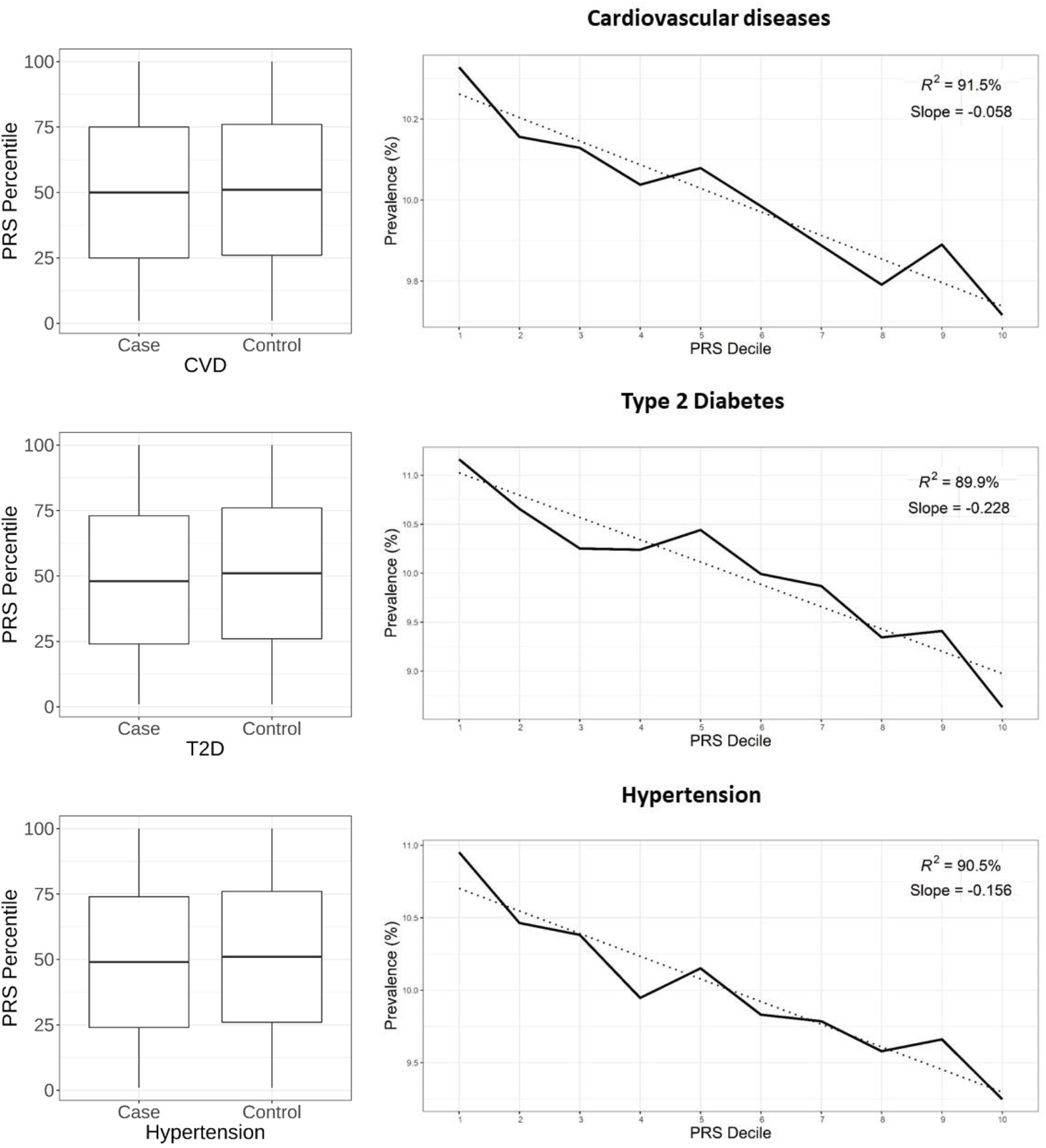
The left panel shows the physical activity PRS percentiles of the cases versus the controls in the FinnGen cohort (n=218,792). Within each boxplot, the horizontal lines reflect the median; the top and bottom of each box reflect the interquartile range; and the whiskers reflect the maximum and minimum values within each grouping. The right panel shows the disease prevalence for cardiovascular diseases (CVD), type 2 diabetes (T2D), and hypertension according to participants physical activity polygenic risk score deciles. High PRS scores denote those genetically predisposed to high physical activity. The dotted line is the trend line, and R^2^ is from the bivariate decile mean–decile model to illustrate the trend linearity between the PRS decile means and the prevalence of the different diseases.

For all three disease types, a higher PA PRS was associated with a lower disease incidence rate (Figure 3, Supplement 1). Participants in the top PA PRS decile had lower odds for developing CVD (OR 0.92, p=2.0*10^−7^), type 2 diabetes (OR=0.81, p=1.2*10^−19^), and hypertension (OR=0.87, p=9.8*10^−15^) (Table 4). In contrast, participants who belonged to the lowest PA PRS percentile had a higher incidence for all three common cardiometabolic diseases after 40 years of age.

**Figure 3.**
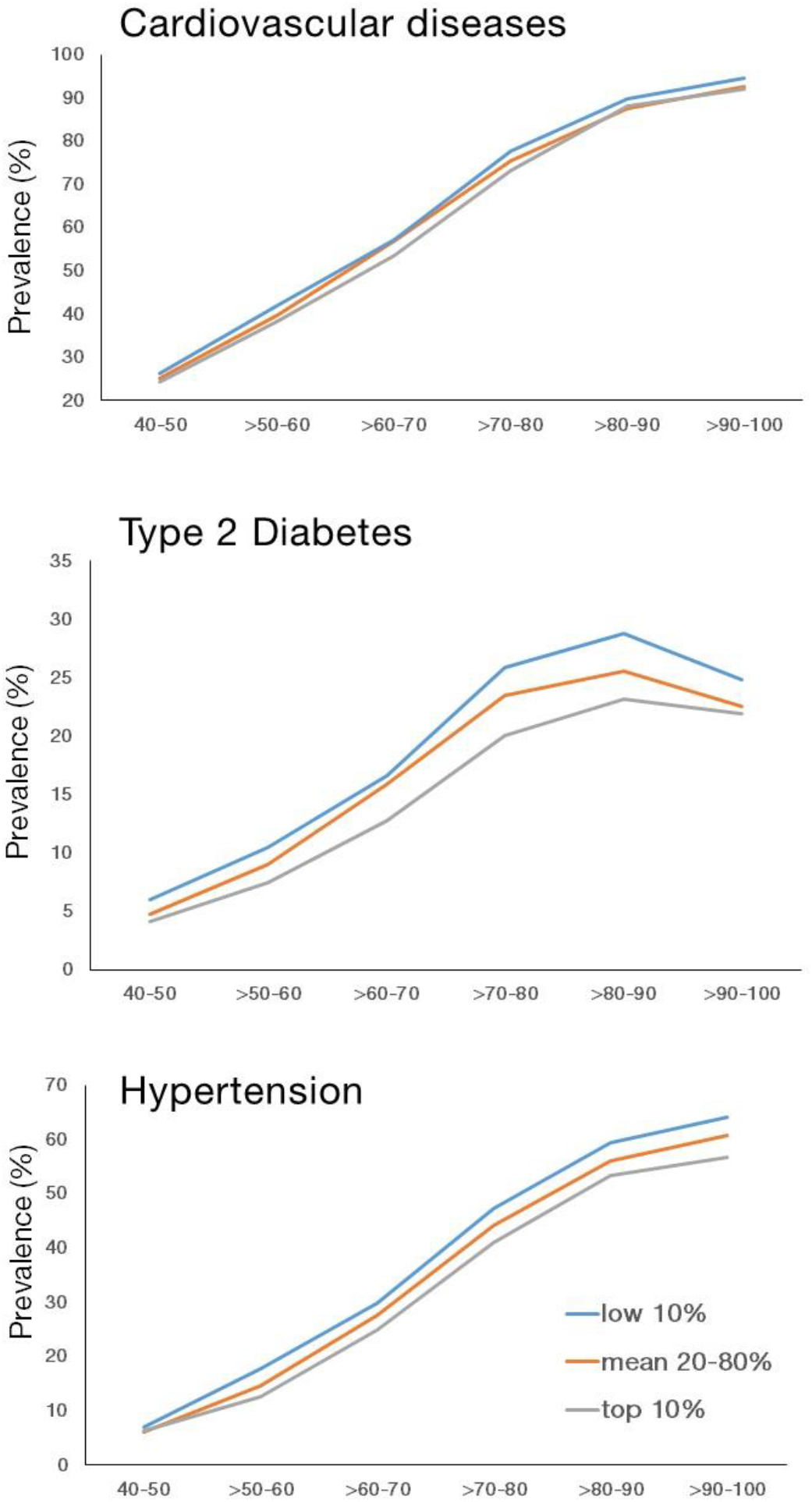
Cumulative disease rate for cardiovascular diseases, type 2 diabetes, and hypertension according to age group and physical activity polygenic risk score categories in the FinnGen cohort (n=218,792).

## DISCUSSION

We adapted a polygenic score for device-based overall physical activity volume [7] and showed that genetically less physically active persons are at higher risk of developing several cardiometabolic diseases and phenotypes when compared to persons with a genetic predisposition for high physical activity. Furthermore, the risk of all-cause mortality was higher among genetically less active individuals. Our results suggest that genetic pleiotropy, i.e., the same genes affecting both physical activity behaviour and CMD risk, may partly explain the associations between low physical activity and higher disease and mortality risk that have frequently reported in literature. In addition, possibly because genetically active persons tend to live longer, it was observed that they are at higher risk of developing Alzheimer’s disease.

There is a substantial amount of literature on the associations between higher physical activity and a lower risk of common CMDs. Moreover, physical activity has been reported to reduce disease risk in a dose-response manner [18]. However, adjusting for genetic confounders in these designs has been impossible except for in twin studies. Physical activity is a multifactorial behavioural and physiological trait influenced by hundreds—if not thousands of genes—that exhibits variation due to hundreds of thousands of genetic variants, most of which are single-nucleotide variants. To our knowledge, this is the first study based on genomic evidence that suggests that a genetic predisposition for physical activity is associated with several cardiometabolic traits. The results are in line with a few twin studies, which have shown that genetic inheritance strongly mediates the association between physical activity and disease risk and mortality [19, 20], as well as a Mendelian randomization study suggesting a bidirectional relationship with physical activity and adiposity [11].

The PA PRS, derived from over million SNPs, does not reflect a genetic predisposition to single underlying mechanism but rather the combined influence of multiple pathways. Genetic risk can lead to activity behaviour through multiple distinct social-behavioural factors [21]. In addition, biological determinants may also play a significant role in regulating physical activity levels [22]. These determinants include three main components—the brain, the cardiorespiratory system, and the muscles—all of which interact within the human body. The optimal function of these organs and systems that participate in energy production predict higher aerobic capacity among individuals representing all age groups. Aerobic capacity is a highly heritable phenotype according to twin studies [15]. In contrast, many pathological conditions related to these functions, such as impaired heart and liver function, vascular structure, and circulatory and metabolic systems, encompass in CMD. It is therefore reasonable to suggest that shared genetic factors that regulate both physical activity and CMD may have a physiological basis.

Recent studies using newly discovered genetic variants for CMD as well as novel methods of generating PRS that use genome-wide variation rather than only a handful of genome-wide significant variants have shown the polygenic nature of CMD and better performance for predicting disease onset [5]. Improved risk prediction over commonly used clinical estimates has been especially found among high-risk individuals representing genetic extremities [23]. The role that PRSs will play in clinical care are currently unclear, but it have been constantly suggested that lifestyle interventions, including physical activity, might be one treatment option for high-risk CMD individuals [23]. Yet, it is not known how high-risk individuals accept and respond to life style treatment. In terms of BMI, it has been shown that the effects of an unhealthy diet, physical activity, and sedentary behaviour on BMI are pronounced in those with a genetic predisposition for a high BMI [24, 25]. We observed here that the prevalence of the three major CMDs (cardiovascular diseases, Type 2 diabetes and hypertension) increased linearly with decreasing physical activity PRS deciles and that the lowest percentiles of genetically less active individuals were at high risk for developing CMDs. We hypothesise, that it is likely that individuals who have high genetic or clinical risk for CMD may also have a “low activity genotype”. This suggest that it can be challenging to intervene with formerly inactive individuals to prevent cardio-metabolic diseases that typically develop over decades. Also exercise training intervention effects on intermediate CMD risk factors are highly individual and suggested to originate from genetic diversities [26, 27]. Future studies investigating lifestyle interventions among individuals representing genetic extremities may reveal how genetic inheritance affects intervention responses or lifestyle modifications.

In addition to its use in CMD, physical activity is also widely recommended as part of the treatment of other diseases. For instance, physical activity has beneficial effects on patients suffering from osteopenia and osteoporosis. In terms of osteoarthrosis, it is known that a physically active lifestyle may either reduce one’s risk of osteoarthrosis through controlling their body weight and improving joint and cartilage structures, but, in particular, sports-related injuries and heavy work-related loading are also known to be predictors of osteoarthritis [28]. This probably explains the lack of an overall association between physical activity PRS and osteoarthrosis in our study. Our results suggest that the genetic inheritance of physical activity is not associated with osteoporosis; thus, the previously reported findings that physical activity has positive effects on bone mineral content and bone strength seem to result from causal mechanistic pathways and not shared genetic effects.

In our study, a higher physical activity PRS, which suggests a genetically active genotype, was associated with a higher risk for developing Alzheimer”s disease. This association was evident especially in atypical or mixed Alzheimer”s diseases diagnosis, while associations with all dementia diagnoses or vascular dementia were not found. In general, physical activity has typically been shown to reduce the risk of Alzheimer”s disease and its brain-related complications [29]. A healthy lifestyle has found to be associated with lower dementia risk—independent of one”s genetic risk level [30]. The risk of Alzheimer”s disease increases strongly with increasing age, but it has been probably overall decreasing in recent years as the overall health of the elderly population has improved globally [31]. In the Finnish population, 15–20% of those over 85 years of age have received an Alzheimer”s diagnosis. We suggest that the association between physical activity PRS and Alzheimer”s disease may partially be explained by improved survival of genetically physically active individuals due to lower risk of early-onset cardiovascular death. However, a formal competing-risk analysis is needed to demonstrate whether this association is robust. Another potential explanation to our findings about increased Alzheimer risk among genetically active individuals is genetic overlap of physical activity and neurodegenerative diseases, previously reported by Doherty et al. [11]. The authors hypothesized that the association might be partially mediated by obesity traits.

Thus far, PRSs have been established for various traits and diseases, but their development for behavioural traits has been limited [32, 33]. Moreover, earlier studies have not assessed how behavioural PRSs predict future major health events—except with regard to alcohol consumption [34]. In this study we investigated associations between the polygenic score for physical activity and multiple register-derived disease endpoints in a large population-based sample (n=218,792). The Finnish personal identity numbers, unique for each individual and utilized in all national digital health care registers, enabled linking the genotyped FinnGen study participants into different health registers. This, in turn, allowed us to test the physical activity PRS with multiple validated ICD-endpoints simultaneously. Our study provides novel information regarding associations between physical activity and common diseases and contributes to the interpretation of sports and exercise science studies.

The polygenic score for physical activity utilized in this study was derived using the UK biobank population representing European ancestry. UK biobank subjects are volunteers and somewhat healthier compared to the general population [35]. However, in the original UK biobank GWAS data, there were many individuals with chronic diseases who were not excluded when constructing the physical activity PRS. It is known that chronic diseases are a common reason for one’s reduction in physical activity [36]; thus, the constructed physical activity PRS may include genetic variants that are primarily predictors of chronic diseases. This needs to be taken into account when interpreting our results. Physical activity PRS was validated utilizing Finnish cohorts [7], and our current association analyses were performed among Finnish citizens. In general, Finns differ from other Europeans primarily in terms of the frequency of less common and rare variants due to genetic isolation and bottlenecks [37]. This different genetic ancestry may limit the generalizability of the physical activity PRS to Finns. Although the physical activity PRS was strongly associated with cardiometabolic traits in our study, the UK Biobank data used to derive PRS for physical activity may have resulted in an underestimation of the associations between PRS for physical activity and cardiometabolic diseases. In contrast, a fraction of FinnGen individuals were collected through hospital biobanks, which may lead to the overestimation of risk [5]. The PRS for physical activity and its associations with CMD need to also be tested in non-European samples.

Today, PRSs for common diseases and lifestyle behaviour can be simultaneously and inexpensively calculated for individuals at birth. The usefulness as well as potential harms of this knowledge needs to be carefully considered before this information is routinely used in health care, i.e., for screening subjects who are at an extreme risk of developing a disease. It is important to consider how absolute and relative risks are assessed and communicated at different stages of life. In addition, there is a need for studies investigating the potential genetic overlap between disease risk and health behaviour in longitudinal settings as well as to increase knowledge regarding how potential interventions and treatments work depending on individuals’ polygenic risk. Like any other genetic risk information, polygenic risk scores are not deterministic. This is demonstrated by studies of monozygotic twins, who have the same genomic sequence and hence identical polygenic scores for any trait or disease. Yet, even the identical twin pairs are more discordant than concordant for all common diseases, indicating the roles of the environment, life style and chance.

### Conclusions

In conclusion, in this study, a higher physical activity PRS was associated with a lower risk for several cardiometabolic diseases and all-cause mortality. These findings highlight the fact that shared genetic factors may modify both health-related behaviour, such as physical activity, and disease risk. The practical applications of polygenic risk information in disease screening as well as for guiding lifestyle and medical interventions remain to be investigated in further studies. An understanding of the individuals’ genetic predisposition to diseases as well as lifestyle factors may help destigmatize individuals who cannot fulfil public lifestyle recommendations.

## METHODS

### Study sample and endpoints

The data comprised 218,792 Finnish citizens from FinnGen, Data Freeze 5. The sample included 56.5% women, and the mean age was 59.8 years (range 1.5 to 120.3). FinnGen includes prospective epidemiological cohorts, diseases-based cohorts, and hospital biobank samples (Data Freeze 5, Supplement 2). In FinnGen, genome information is combined with national hospital discharge (1968–present), death (1969-present), cancer (1953–present), and the Social insurance Institute of Finland (Kela) medication reimbursement (1995–present) registers. Endpoint definitions were based on International Statistical Classification of Diseases and Related Health Problems (ICD-8, ICD-9 and ICD-10) codes. The ICD-codes included in each endpoint can be revised at FinnGen webpages (https://www.finngen.fi/en/researchers/clinical-endpoints, DF-5). The quality of the CMD diagnoses in these registers has been extensive validated in several studies [38]. For example, health care data included 21,012 major coronary heart disease events, 55,970 hypertension cases, and 29,139 T2D cases (Tables 1–3). We also tested associations with PA PRS and CMD medication endpoints. Based on the three existing smoking status variables in FinnGen, 40.2% (n=87,859) had missing smoking status data, while 59.8% (n=130,933) could be classified. Of the latter, 22.9% (29,961) were current smokers, 24% (31471) former smokers, 51.1% (66,872) were never smokers and 2.0% (2629) were non-current smokers (it was not known whether they were former smokers or never smokers). Occasional smokers were considered current smokers. Based on this information, we decided to use data from current smokers and never smokers in our analysis.

### Genotyping, quality control, and imputation

The FinnGen Study samples were genotyped with various Illumina and custom AxiomGT1 Affymetrix arrays (Thermo Fisher Scientific, Santa Clara, CA, USA; please see http://www.finngen.fi/en/researchers/genotyping and Supplement 3.

### Polygenic scoring for physical activity

The PA PRS, which was recently developed for continuous accelerometer-based overall PA volume [7, 39] was adapted to the FinnGen cohort. Briefly, GWAS summary statistics from the UK Biobank for risk score calculation were obtained from the data sharing repository of the GWAS of PA measured by an accelerometer [11]. The objective assessment of PA was measured for a seven-day period using an Axivity AX3 wrist-worn triaxial accelerometer in the UK Biobank cohort (n=103,702). The non-wear time was detected and imputed by the expert working group, resulting in a total PA calculated by averaging all worn and imputed values [11, 39]. To obtain PRSs for PA, we used a Bayesian approach, accounted for linkage disequilibrium (LDPred) [40], and adjusted for the LD reference panel of unrelated Finnish individuals from the national FINRISK study (n=27,284) [17]. The total number of variants used for risk score calculation in our first analyses was 1,140,182.

### Statistical analyses

Associations between PA PRS and body mass index (BMI), common diseases, and mortality were analysed with linear and logistic regression models adjusted for age, gender, and the 10 principal components of ancestry. An increase in risk was calculated per 1 SD change in PRS. The distribution of four SDs covered 95% of the population. The number of disease endpoints varied between 894 (non-alcoholic fatty level disease) and 111,108 (cardiovascular diseases, Tables 1–3). The false discovery rate (FDR) was used to correct the p-values for multiple testing [41], and the significance threshold was set to p?<?0.05. Three major disease endpoints with highly significant associations with PA PRS were selected for more detailed analyses (cardiovascular diseases p=9.5*10^−19^, Type 2 diabetes p=4.1*10^−42^, and hypertension p=2.7*10^−44^). Individuals were binned into three categories according to their PRS decile (top 10%, 20–80%, low 10%), and the unadjusted prevalence of disease within each bin was determined. The proportion of the cohort and diseased individuals with a specific polygenic risk level was determined by comparing progressively more extreme tails of the distribution with the remainder of the population in a logistic regression model predicting the disease endpoint and adjusted for age, gender, and the 10 principal components of ancestry.

### Ethical statements

The patients and control subjects in FinnGen provided their informed consent for biobank research based on the Finnish Biobank Act. Alternatively, older research cohorts, collected prior the start of FinnGen (in August 2017), were collected based on study-specific consent and later transferred to the Finnish biobanks after approval by Fimea, the National Supervisory Authority for Welfare and Health. The recruitment protocols followed the biobank protocols approved by Fimea. The Coordinating Ethics Committee of the Hospital District of Helsinki and Uusimaa (HUS) approved the FinnGen study protocol Nr HUS/990/2017.

The FinnGen study is approved by the Finnish Institute for Health and Welfare (permit numbers: THL/2031/6.02.00/2017, THL/1101/5.05.00/2017, THL/341/6.02.00/2018, THL/2222/6.02.00/2018, THL/283/6.02.00/2019, THL/1721/5.05.00/2019, THL/1524/5.05.00/2020, and THL/2364/14.02/2020), Digital and Population Data Service Agency (permit numbers: VRK43431/2017-3, VRK/6909/2018-3, VRK/4415/2019-3), the Social Insurance Institution (permit numbers: Kela 58/522/2017, Kela 131/522/2018, Kela 70/522/2019, Kela 98/522/2019, Kela 138/522/2019, Kela 2/522/2020, Kela 16/522/2020), and Statistics Finland (permit numbers: TK-53-1041-17 and TK-53-90-20).

The Biobank Access Decisions for FinnGen samples and the data utilized in FinnGen Data Freeze 6 include: THL Biobank BB2017_55, BB2017_111, BB2018_19, BB_2018_34, BB_2018_67, BB2018_71, BB2019_7, BB2019_8, BB2019_26, BB2020_1, Finnish Red Cross Blood Service Biobank 7.12.2017, Helsinki Biobank HUS/359/2017, Auria Biobank AB17-5154, Biobank Borealis of Northern Finland_2017_1013, Biobank of Eastern Finland 1186/2018, Finnish Clinical Biobank Tampere MH0004, Central Finland Biobank 1-2017, and Terveystalo Biobank STB 2018001.

## Data Availability

The FinnGen data may be accessed through Finnish Biobanks FinBB portal.

https://www.finbb.fi/

## Acknowledgments

The FinnGen project is funded by two grants from Business Finland (HUS 4685/31/2016 and UH 4386/31/2016) and the following industry partners: AbbVie Inc., AstraZeneca UK Ltd, Biogen MA Inc., Celgene Corporation, Celgene International II Sàrl, Genentech Inc., Merck Sharp & Dohme Corp, Pfizer Inc., GlaxoSmithKline Intellectual Property Development Ltd., Sanofi US Services Inc., Maze Therapeutics Inc., Janssen Biotech Inc, and Novartis AG. Following biobanks are acknowledged for the project samples: Auria Biobank (www.auria.fi/biopankki), THL Biobank (www.thl.fi/biobank), Helsinki Biobank (www.helsinginbiopankki.fi), Biobank Borealis of Northern Finland (https://www.ppshp.fi/Tutkimus-ja-opetus/Biopankki/Pages/Biobank-Borealis-briefly-in-English.aspx), Finnish Clinical Biobank Tampere (www.tays.fi/en-US/Research_and_development/Finnish_Clinical_Biobank_Tampere), Biobank of Eastern Finland (www.ita-suomenbiopankki.fi/en), Central Finland Biobank (www.ksshp.fi/fi-FI/Potilaalle/Biopankki), Finnish Red Cross Blood Service Biobank (www.veripalvelu.fi/verenluovutus/biopankkitoiminta), and Terveystalo Biobank (www.terveystalo.com/fi/Yritystietoa/Terveystalo-Biopankki/Biopankki/). All Finnish Biobanks are members of BBMRI.fi infrastructure (www.bbmri.fi) and FinBB (https://finbb.fi/).

ES was supported by the Juho Vainio Foundation and the Päivikki and Sakari Sohlberg Foundation. SR was supported by the Academy of the Finland Center of Excellence in Complex Disease Genetics (Grant No 312062 and 336820), the Finnish Foundation for Cardiovascular Research, the Sigrid Juselius Foundation, and the University of Helsinki HiLIFE Fellow and Grand Challenge grants.

The funders had no roles in the study design, collection, analysis, or interpretations of the data or in the publication process. The authors have no conflicts of interest to report. All authors declare that the results of the study are presented clearly, honestly, and without fabrication, falsification, or inappropriate data manipulation. We wish to thank all affiliated biobanks and the FinnGen participants for their valuable contribution to science.

## Author contributions

E.S., U.K. and J.K. conceived the idea and applied permission to use FinnGen data for the current plan. J.K., U.K., S.R. directed and supervised the study. T.P. and E.S. planned the analysis and T.P. performed the analyses. E.S. drafted the first version of manuscript. T.P., U.K. and J.K. provided critical feedback and key elements in interpreting the results. All authors contributed to editing and approval of the final paper.

## List of Supplemental Digital Content

Supplemental Digital Content 1; Table 1. Cumulative disease rate according to age group and physical activity polygenic risk score category in FinnGen cohort (n=218,792). Supplemental Digital Content 2; List of FinnGen data Freeze 5 cohorts. Supplemental Digital Content 3; Genotyping and quality control of the FinnGen data. Supplemental Digital Content 4; Contributors of FinnGen.

