## Supplementary files for "Polygenic score for physical activity provides odds for multiple common diseases"

Supplementary file 1. Table 1. Cumulative disease rate according to the age group and physical activity polygenic risk score category in FinnGen cohort (N=218,792).

| Disease | Polygenic score category | Age group | N cases/controls | Cumulative% |
|---|---|---|---|---|
| Cardiovascular diseases | low 10% | 40-50 | 632/1773 | 26 |
| | | >50-60 | 1583/2198 | 42 |
| | | >60-70 | 2766/2070 | 57 |
| | | >70-80 | 3739/1074 | 78 |
| | | >80-90 | 1894/219 | 90 |
| | | >90-100 | 319/18 | 95 |
| | mean 20 to 80% | 40-50 | 4973/14812 | 25 |
| | | >50-60 | 11979/18094 | 40 |
| | | >60-70 | 22207/16912 | 57 |
| | | >70-80 | 28191/9174 | 75 |
| | | >80-90 | 14910/2117 | 88 |
| | | >90-100 | 2423/198 | 92 |
| | top 10% | 40-50 | 593/1837 | 24 |
| | | >50-60 | 1417/2271 | 38 |
| | | >60-70 | 2646/2288 | 54 |
| | | >70-80 | 3421/1247 | 73 |
| | | >80-90 | 1886/255 | 88 |
| | | >90-100 | 320/28 | 92 |
| Type 2 Diabetes | low 10% | 40-50 | 137/2148 | 6 |
| | | >50-60 | 386/3284 | 11 |
| | | >60-70 | 778/3908 | 17 |
| | | >70-80 | 1209/3475 | 26 |
| | | >80-90 | 594/1468 | 29 |
| | | >90-100 | 82/248 | 25 |
| | mean 20 to 80% | 40-50 | 890/18002 | 5 |
| | | >50-60 | 2638/26693 | 9 |
| | | >60-70 | 6072/32048 | 16 |
| | | >70-80 | 8556/27907 | 23 |
| | | >80-90 | 4265/12420 | 26 |
| | | >90-100 | 582/2001 | 23 |
| | top 10% | 40-50 | 95/2220 | 4 |
| | | >50-60 | 271/3337 | 8 |
| | | >60-70 | 614/4198 | 13 |
| | | >70-80 | 921/3667 | 20 |
| | | >80-90 | 484/1606 | 23 |
| | | >90-100 | 75/268 | 22 |
| Hypertension | low 10% | 40-50 | 167/2238 | 7 |
| | | >50-60 | 672/3109 | 18 |
| | | >60-70 | 1447/3389 | 30 |
| | | >70-80 | 2280/2533 | 47 |
| | | >80-90 | 1254/859 | 59 |
| | | >90-100 | 216/121 | 64 |
| | mean 20 to 80% | 40-50 | 1215/18570 | 6 |
| | | >50-60 | 4385/25688 | 15 |
| | | >60-70 | 10762/28357 | 28 |
| | | >70-80 | 16532/20833 | 44 |
| | | >80-90 | 9532/7495 | 56 |
| | | >90-100 | 1591/1030 | 61 |
| | top 10% | 40-50 | 151/2279 | 6 |
| | | >50-60 | 460/3228 | 12 |
| | | >60-70 | 1231/3703 | 25 |
| | | >70-80 | 1917/2751 | 41 |
| | | >80-90 | 1140/1001 | 53 |
| | | >90-100 | 197/151 | 57 |

Supplementary file 2.

**FinnGen data freeze 5 cohorts**

AURIA BIOBANK

BOREALIS BIOBANK

HELSINKI BIOBANK

BIOBANK OF CENTRAL FINLAND

BIOBANK OF EASTERN FINLAND

FINNISH CLINICAL BIOBANK TAMPERE

BLOOD SERVICE BIOBANK

TERVEYSTALO BB

THL BB:

BOTNIA

COROGENE

FINRISK 1992-2012

FINHEALTH 2017

FinIPF

GENERISK

HEALTH 2000/H2011

HHS

KUUSAMO (=FR11)

MIGRAINE

SUPER

T1D

TWINS

Supplementary file 3.

**Genotyping and quality control of the FinnGen data**

Chip genotyping were done using several Illumina and Affymetrix FinnGen Axiom arrays. The algorithm for genotype calling were GenCall or GenCall+zCall for Illumina and AxiomGT1 for Affymetrix chip genotypes. The genome build of all genotypes were set to GRCh38/hg38. Genotype quality control were done in batch-wise based on array manufacturer and version. From all genotyping batches, variants with call rate below 98%, minor allele count below 3 and Hardy-Weinberg Equilibrium p-value lower than 1e-06 were removed. Also samples from all batches with call rate below 95% and heterozygosity test method-of-moments F coefficient estimate value deviated more than ±4SD from the mean were removed along with the samples which failed sex check or were among the multi-dimensional scaling principal component analysis outliers.

Pre-phasing were performed using Eagle v2.3 [1] and imputation with Beagle v4.1 [2]. Genotypes of all batches were imputed to Sequencing Initiative Suomi v3 reference panel which consist of 3775 Finnish whole genome sequences (depth up to 30x) having total of 16962023 variants. As post-imputation quality control, variants with imputation quality score below 0.7 were removed.

1 : Loh et al. Reference-based phasing using the Haplotype Reference Consortium panel. Nature Genetics 2016.

2: Browning et al. Genotype imputation with millions of reference samples. American Journal of Human Genetics 2016.

Supplementary file 4.

### Contributors of FinnGen

#### Steering Committee

Aarno Palotie Institute for Molecular Medicine Finland, HiLIFE, University of Helsinki, Finland
Mark Daly Institute for Molecular Medicine Finland, HiLIFE, University of Helsinki, Finland

**Pharmaceutical companies**

Bridget Riley-Gills Abbvie, Chicago, IL, United States
Howard Jacob Abbvie, Chicago, IL, United States
Dirk Paul Astra Zeneca, Cambridge, United Kingdom
Heiko Runz Biogen, Cambridge, MA, United States
Sally John Biogen, Cambridge, MA, United States
Robert Plenge Celgene, Summit, NJ, United States/Bristol Myers Squibb, New York, NY, United States
Mark McCarthy Genentech, San Francisco, CA, United States
Julie Hunkapiller Genentech, San Francisco, CA, United States
Meg Ehm GlaxoSmithKline, Brentford, United Kingdom
Caroline Fox Merck, Kenilworth, NJ, United States
Anders Mälarstig Pfizer, New York, NY, United States
Katherine Klinger Sanofi, Paris, France
Katherine Call Sanofi, Paris, France
Tim Behrens Maze Therapeutics, San Francisco, CA, United States
Robert Yang Janssen Biotech, Beerse, Belgium
Richard Siegel Novartis, Basel, Switzerland

**University of Helsinki & Biobanks**

Tomi Mäkelä HiLIFE, University of Helsinki, Finland, Finland
Jaakko Kaprio Institute for Molecular Medicine Finland, HiLIFE, Helsinki, Finland, Finland
Petri Virolainen Auria Biobank / University of Turku / Hospital District of Southwest Finland, Turku, Finland
Antti Hakanen Auria Biobank / University of Turku / Hospital District of Southwest Finland, Turku, Finland
Terhi Kilpi THL Biobank / The National Institute of Health and Welfare Helsinki, Finland
Markus Perola THL Biobank / The National Institute of Health and Welfare Helsinki, Finland
Jukka Partanen Finnish Red Cross Blood Service / Finnish Hematology Registry and Clinical Biobank, Helsinki, Finland
Anne Pitkäranta Helsinki Biobank / Helsinki University and Hospital District of Helsinki and Uusimaa, Helsinki
Juhani Junttila Northern Finland Biobank Borealis / University of Oulu / Northern Ostrobothnia Hospital District, Oulu, Finland
Raisa Serpi Northern Finland Biobank Borealis / University of Oulu / Northern Ostrobothnia Hospital District, Oulu, Finland
Tarja Laitinen Finnish Clinical Biobank Tampere / University of Tampere / Pirkanmaa Hospital District, Tampere, Finland
Johanna Mäkelä Finnish Clinical Biobank Tampere / University of Tampere / Pirkanmaa Hospital District, Tampere, Finland
Veli-Matti Kosma Biobank of Eastern Finland / University of Eastern Finland / Northern Savo Hospital District, Kuopio, Finland
Urho Kujala Central Finland Biobank / University of Jyväskylä / Central Finland Health Care District, Jyväskylä, Finland

**Other Experts/ Non-Voting Members**

Outi Tuovila Business Finland, Helsinki, Finland
Raimo Pakkanen Business Finland, Helsinki, Finland

### Scientific Committee

**Pharmaceutical companies**


Jeffrey Waring Abbvie, Chicago, IL, United States
Ali Abbasi Abbvie, Chicago, IL, United States
Mengzhen Liu Abbvie, Chicago, IL, United States
Ioanna Tachmazidou Astra Zeneca, Cambridge, United Kingdom
Chia-Yen Chen Biogen, Cambridge, MA, United States
Heiko Runz Biogen, Cambridge, MA, United States
Shameek Biswas Celgene, Summit, NJ, United States/Bristol Myers Squibb, New York, NY, United States
Julie Hunkapiller Genentech, San Francisco, CA, United States
Meg Ehm GlaxoSmithKline, Brentford, United Kingdom
Neha Raghavan Merck, Kenilworth, NJ, United States
Aparna Chhibber Merck, Kenilworth, NJ, United States
Anders Mälarstig Pfizer, New York, NY, United States
Xinli Hu Pfizer, New York, NY, United States
Katherine Call Sanofi, Paris, France
Katherine Klinger Sanofi, Paris, France
Matthias Gossel Sanofi, Paris, France
Robert Graham Maze Therapeutics, San Francisco, CA, United States
Tim Behrens Maze Therapeutics, San Francisco, CA, United States
Beryl Cummings Maze Therapeutics, San Francisco, CA, United States
Wilco Fleuren Janssen Biotech, Beerse, Belgium
Dawn Waterworth Janssen Biotech, Beerse, Belgium
Nicole Renaud Novartis, Basel, Switzerland
Aviv Madar Novartis, Basel, Switzerland
Maen Obeidat Novartis, Basel, Switzerland


**University of Helsinki & Biobanks**


Samuli Ripatti Institute for Molecular Medicine Finland, HiLIFE, Helsinki, Finland
Johanna Schleutker Auria Biobank / Univ. of Turku / Hospital District of Southwest Finland, Turku, Finland
Markus Perola THL Biobank / The National Institute of Health and Welfare Helsinki, Finland
Mikko Arvas Finnish Red Cross Blood Service / Finnish Hematology Registry and Clinical Biobank, Helsinki, Finland
Olli Carpén Helsinki Biobank / Helsinki University and Hospital District of Helsinki and Uusimaa, Helsinki
Reetta Hinttala Northern Finland Biobank Borealis / University of Oulu / Northern Ostrobothnia Hospital District, Oulu, Finland
Johannes Kettunen Northern Finland Biobank Borealis / University of Oulu / Northern Ostrobothnia Hospital District, Oulu, Finland
Johanna Mäkelä Finnish Clinical Biobank Tampere / University of Tampere / Pirkanmaa Hospital District, Tampere, Finland
Arto Mannermaa Biobank of Eastern Finland / University of Eastern Finland / Northern Savo Hospital District, Kuopio, Finland
Jari Laukkanen Central Finland Biobank / University of Jyväskylä / Central Finland Health Care District, Jyväskylä, Finland
Urho Kujala Central Finland Biobank / University of Jyväskylä / Central Finland Health Care District, Jyväskylä, Finland


### Clinical Groups

**Neurology Group**


Reetta Kälviäinen Northern Savo Hospital District, Kuopio, Finland
Valtteri Julkunen Northern Savo Hospital District, Kuopio, Finland
Hilkka Soininen Northern Savo Hospital District, Kuopio, Finland
Anne Remes Northern Ostrobothnia Hospital District, Oulu, Finland
Mikko Hiltunen Northern Savo Hospital District, Kuopio, Finland

| | |
|---|---|
| Jukka Peltola | Pirkanmaa Hospital District, Tampere, Finland |
| Pentti Tienari | Hospital District of Helsinki and Uusimaa, Helsinki, Finland |
| Juha Rinne | Hospital District of Southwest Finland, Turku, Finland |
| Roosa Kallionpää | Hospital District of Southwest Finland, Turku, Finland |
| Ali Abbasi | Abbvie, Chicago, IL, United States |
| Adam Ziemann | Abbvie, Chicago, IL, United States |
| Jeffrey Waring | Abbvie, Chicago, IL, United States |
| Sahar Esmaeeli | Abbvie, Chicago, IL, United States |
| Nizar Smaoui | Abbvie, Chicago, IL, United States |
| Anne Lehtonen | Abbvie, Chicago, IL, United States |
| Susan Eaton | Biogen, Cambridge, MA, United States |
| Heiko Runz | Biogen, Cambridge, MA, United States |
| Sanni Lahdenperä | Biogen, Cambridge, MA, United States |
| Janet van Adelsberg | Celgene, Summit, NJ, United States/ Bristol Myers Squibb, New York, NY, United States |
| Shameek Biswas | Celgene, Summit, NJ, United States/ Bristol Myers Squibb, New York, NY, United States |
| Julie Hunkapiller | Genentech, San Francisco, CA, United States |
| Natalie Bowers | Genentech, San Francisco, CA, United States |
| Edmond Teng | Genentech, San Francisco, CA, United States |
| Sarah Pendergrass | Genentech, San Francisco, CA, United States |
| Onuralp Soylemez | Merck, Kenilworth, NJ, United States |
| Kari Linden | Pfizer, New York, NY, United States |
| Fanli Xu | GlaxoSmithKline, Brentford, United Kingdom |
| David Pulford | GlaxoSmithKline, Brentford, United Kingdom |
| Kirsi Auro | GlaxoSmithKline, Brentford, United Kingdom |
| Laura Addis | GlaxoSmithKline, Brentford, United Kingdom |
| John Eicher | GlaxoSmithKline, Brentford, United Kingdom |
| Minna Raivio | Hospital District of Helsinki and Uusimaa, Helsinki, Finland |
| Sarah Pendergrass | Genentech, San Francisco, CA, United States |
| Beryl Cummings | Maze Therapeutics, San Francisco, CA, United States |
| Juulia Partanen | Institute for Molecular Medicine Finland, HiLIFE, University of Helsinki, Finland |

**Gastroenterology Group**

| | |
|---|---|
| Martti Färkkilä | Hospital District of Helsinki and Uusimaa, Helsinki, Finland |
| Jukka Koskela | Hospital District of Helsinki and Uusimaa, Helsinki, Finland |
| Sampsa Pikkarainen | Hospital District of Helsinki and Uusimaa, Helsinki, Finland |
| Airi Jussila | Pirkanmaa Hospital District, Tampere, Finland |
| Katri Kaukinen | Pirkanmaa Hospital District, Tampere, Finland |
| Timo Blomster | Northern Ostrobothnia Hospital District, Oulu, Finland |
| Mikko Kiviniemi | Northern Savo Hospital District, Kuopio, Finland |
| Markku Voutilainen | Hospital District of Southwest Finland, Turku, Finland |
| Ali Abbasi | Abbvie, Chicago, IL, United States |
| Graham Heap | Abbvie, Chicago, IL, United States |
| Jeffrey Waring | Abbvie, Chicago, IL, United States |
| Nizar Smaoui | Abbvie, Chicago, IL, United States |
| Fedik Rahimov | Abbvie, Chicago, IL, United States |
| Anne Lehtonen | Abbvie, Chicago, IL, United States |
| Keith Usiskin | Celgene, Summit, NJ, United States/ Bristol Myers Squibb, New York, NY, United States |
| Tim Lu | Genentech, San Francisco, CA, United States |
| Natalie Bowers | Genentech, San Francisco, CA, United States |
| Danny Oh | Genentech, San Francisco, CA, United States |
| Sarah Pendergrass | Genentech, San Francisco, CA, United States |
| Kirsi Kalpala | Pfizer, New York, NY, United States |
| Melissa Miller | Pfizer, New York, NY, United States |
| Xinli Hu | Pfizer, New York, NY, United States |

Linda McCarthy GlaxoSmithKline, Brentford, United Kingdom
Onuralp Soylemez Merck, Kenilworth, NJ, United States
Mark Daly Institute for Molecular Medicine Finland, HiLIFE, University of Helsinki, Finland

**Rheumatology Group**
Kari Eklund Hospital District of Helsinki and Uusimaa, Helsinki, Finland
Antti Palomäki Hospital District of Southwest Finland, Turku, Finland
Pia Isomäki Pirkanmaa Hospital District, Tampere, Finland
Laura Pirilä Hospital District of Southwest Finland, Turku, Finland
Oili Kaipiainen-Seppänen Northern Savo Hospital District, Kuopio, Finland
Johanna Huhtakangas Northern Ostrobothnia Hospital District, Oulu, Finland
Ali Abbasi Abbvie, Chicago, IL, United States
Jeffrey Waring Abbvie, Chicago, IL, United States
Fedik Rahimov Abbvie, Chicago, IL, United States
Apinya Lertratanakul Abbvie, Chicago, IL, United States
Nizar Smaoui Abbvie, Chicago, IL, United States
Anne Lehtonen Abbvie, Chicago, IL, United States
David Close Astra Zeneca, Cambridge, United Kingdom
Marla Hochfeld Celgene, Summit, NJ, United States/ Bristol Myers Squibb, New York, NY, United States
Natalie Bowers Genentech, San Francisco, CA, United States
Sarah Pendergrass Genentech, San Francisco, CA, United States
Onuralp Soylemez Merck, Kenilworth, NJ, United States
Kirsi Kalpala Pfizer, New York, NY, United States
Nan Bing Pfizer, New York, NY, United States
Xinli Hu Pfizer, New York, NY, United States
Jorge Esparza Gordillo GlaxoSmithKline, Brentford, United Kingdom
Kirsi Auro GlaxoSmithKline, Brentford, United Kingdom
Dawn Waterworth Janssen Biotech, Beerse, Belgium
Nina Mars Institute for Molecular Medicine Finland, HiLIFE, Helsinki, Finland

**Pulmonology Group**
Tarja Laitinen Pirkanmaa Hospital District, Tampere, Finland
Margit Pelkonen Northern Savo Hospital District, Kuopio, Finland
Paula Kauppi Hospital District of Helsinki and Uusimaa, Helsinki, Finland
Hannu Kankaanranta Pirkanmaa Hospital District, Tampere, Finland
Terttu Harju Northern Ostrobothnia Hospital District, Oulu, Finland
Riitta Lahesmaa Hospital District of Southwest Finland, Turku, Finland

Nizar Smaoui Abbvie, Chicago, IL, United States
Alex Mackay Astra Zeneca, Cambridge, United Kingdom
Glenda Lassi Astra Zeneca, Cambridge, United Kingdom
Susan Eaton Biogen, Cambridge, MA, United States
Steven Greenberg Celgene, Summit, NJ, United States/ Bristol Myers Squibb, New York, NY, United States
Hubert Chen Genentech, San Francisco, CA, United States
Sarah Pendergrass Genentech, San Francisco, CA, United States
Natalie Bowers Genentech, San Francisco, CA, United States
Joanna Betts GlaxoSmithKline, Brentford, United Kingdom
Soumitra Ghosh GlaxoSmithKline, Brentford, United Kingdom
Kirsi Auro GlaxoSmithKline, Brentford, United Kingdom
Rajashree Mishra GlaxoSmithKline, Brentford, United Kingdom
Sina Rüeger Institute for Molecular Medicine Finland, HiLIFE, University of Helsinki, Finland

**Cardiometabolic Diseases Group**
Teemu Niiranen The National Institute of Health and Welfare Helsinki, Finland
Felix Vaura The National Institute of Health and Welfare Helsinki, Finland

| | |
|---|---|
| Veikko Salomaa | The National Institute of Health and Welfare Helsinki, Finland |
| Markus Juonala | Hospital District of Southwest Finland, Turku, Finland |
| Kaj Metsärinne | Hospital District of Southwest Finland, Turku, Finland |
| Mika Kähönen | Pirkanmaa Hospital District, Tampere, Finland |
| Juhani Junttila | Northern Ostrobothnia Hospital District, Oulu, Finland |
| Markku Laakso | Northern Savo Hospital District, Kuopio, Finland |
| Jussi Pihlajamäki | Northern Savo Hospital District, Kuopio, Finland |
| Daniel Gordin | Hospital District of Helsinki and Uusimaa, Helsinki, Finland |
| Juha Sinisalo | Hospital District of Helsinki and Uusimaa, Helsinki, Finland |
| Marja-Riitta Taskinen | Hospital District of Helsinki and Uusimaa, Helsinki, Finland |
| Tiinamaija Tuomi | Hospital District of Helsinki and Uusimaa, Helsinki, Finland |
| Jari Laukkanen | Central Finland Health Care District, Jyväskylä, Finland |
| Benjamin Challis | Astra Zeneca, Cambridge, United Kingdom |
| Dirk Paul | Astra Zeneca, Cambridge, United Kingdom |
| Julie Hunkapiller | Genentech, San Francisco, CA, United States |
| Natalie Bowers | Genentech, San Francisco, CA, United States |
| Sarah Pendergrass | Genentech, San Francisco, CA, United States |
| Onuralp Soylemez | Merck, Kenilworth, NJ, United States |
| Jaakko Parkkinen | Pfizer, New York, NY, United States |
| Melissa Miller | Pfizer, New York, NY, United States |
| Russell Miller | Pfizer, New York, NY, United States |
| Audrey Chu | GlaxoSmithKline, Brentford, United Kingdom |
| Kirsi Auro | GlaxoSmithKline, Brentford, United Kingdom |
| Keith Usiskin | Celgene, Summit, NJ, United States/ Bristol Myers Squibb, New York, NY, United States |
| Amanda Elliott | Institute for Molecular Medicine Finland, HiLIFE, University of Helsinki, Finland / Broad Institute, Cambridge, MA, United States |
| Joel Rämö | Institute for Molecular Medicine Finland, HiLIFE, University of Helsinki, Finland |
| Samuli Ripatti | Institute for Molecular Medicine Finland, HiLIFE, University of Helsinki, Finland |
| Mary Pat Reeve | Institute for Molecular Medicine Finland, HiLIFE, University of Helsinki, Finland |
| Sanni Ruotsalainen | Institute for Molecular Medicine Finland, HiLIFE, University of Helsinki, Finland |

**Oncology Group**

| | |
|---|---|
| Tuomo Meretoja | Hospital District of Helsinki and Uusimaa, Helsinki, Finland |
| Heikki Joensuu | Hospital District of Helsinki and Uusimaa, Helsinki, Finland |
| Olli Carpén | Hospital District of Helsinki and Uusimaa, Helsinki, Finland |
| Lauri Aaltonen | Hospital District of Helsinki and Uusimaa, Helsinki, Finland |
| Johanna Mattson | Hospital District of Helsinki and Uusimaa, Helsinki, Finland |
| Annika Auranen | Pirkanmaa Hospital District , Tampere, Finland |
| Peeter Karihtala | Northern Ostrobothnia Hospital District, Oulu, Finland |
| Saila Kauppila | Northern Ostrobothnia Hospital District, Oulu, Finland |
| Päivi Auvinen | Northern Savo Hospital District, Kuopio, Finland |
| Klaus Elenius | Hospital District of Southwest Finland, Turku, Finland |
| Johanna Schleutker | Hospital District of Southwest Finland, Turku, Finland |
| Relja Popovic | Abbvie, Chicago, IL, United States |
| Jeffrey Waring | Abbvie, Chicago, IL, United States |
| Bridget Riley-Gillis | Abbvie, Chicago, IL, United States |
| Anne Lehtonen | Abbvie, Chicago, IL, United States |
| Jennifer Schutzman | Genentech, San Francisco, CA, United States |
| Julie Hunkapiller | Genentech, San Francisco, CA, United States |
| Natalie Bowers | Genentech, San Francisco, CA, United States |
| Sarah Pendergrass | Genentech, San Francisco, CA, United States |
| Andrey Loboda | Merck, Kenilworth, NJ, United States |
| Aparna Chhibber | Merck, Kenilworth, NJ, United States |
| Heli Lehtonen | Pfizer, New York, NY, United States |
| Stefan McDonough | Pfizer, New York, NY, United States |

Marika Crohns Sanofi, Paris, France
Sauli Vuoti Sanofi, Paris, France
Diptee Kulkarni GlaxoSmithKline, Brentford, United Kingdom
Kirsi Auro GlaxoSmithKline, Brentford, United Kingdom
Esa Pitkänen Institute for Molecular Medicine Finland, HiLIFE, University of Helsinki, Finland
Nina Mars Institute for Molecular Medicine Finland, HiLIFE, University of Helsinki, Finland
Mark Daly Institute for Molecular Medicine Finland, HiLIFE, University of Helsinki, Finland

**Opthalmology Group**
Kai Kaarniranta Northern Savo Hospital District, Kuopio, Finland
Joni A Turunen Hospital District of Helsinki and Uusimaa, Helsinki, Finland
Terhi Ollila Hospital District of Helsinki and Uusimaa, Helsinki, Finland
Sanna Seitsonen Hospital District of Helsinki and Uusimaa, Helsinki, Finland
Hannu Uusitalo Pirkanmaa Hospital District, Tampere, Finland
Vesa Aaltonen Hospital District of Southwest Finland, Turku, Finland
Hannele Uusitalo-Järvinen Pirkanmaa Hospital District, Tampere, Finland
Marja Luodonpää Northern Ostrobothnia Hospital District, Oulu, Finland
Nina Hautala Northern Ostrobothnia Hospital District, Oulu, Finland
Mengzhen Liu Abbvie, Chicago, IL, United States
Heiko Runz Biogen, Cambridge, MA, United States
Stephanie Loomis Biogen, Cambridge, MA, United States
Erich Strauss Genentech, San Francisco, CA, United States
Natalie Bowers Genentech, San Francisco, CA, United States
Hao Chen Genentech, San Francisco, CA, United States
Sarah Pendergrass Genentech, San Francisco, CA, United States
Anna Podgornaia Merck, Kenilworth, NJ, United States
Juha Karjalainen Institute for Molecular Medicine Finland, HiLIFE, University of Helsinki, Finland / Broad Institute, Cambridge, MA, United States
Esa Pitkänen Institute for Molecular Medicine Finland, HiLIFE, University of Helsinki, Finland

**Dermatology Group**
Kaisa Tasanen Northern Ostrobothnia Hospital District, Oulu, Finland
Laura Huilaja Northern Ostrobothnia Hospital District, Oulu, Finland
Katariina Hannula-Jouppi Hospital District of Helsinki and Uusimaa, Helsinki, Finland
Teea Salmi Pirkanmaa Hospital District, Tampere, Finland
Sirkku Peltonen Hospital District of Southwest Finland, Turku, Finland
Leena Koulu Hospital District of Southwest Finland, Turku, Finland

Kirsi Kalpala Pfizer, New York, NY, United States
Ying Wu Pfizer, New York, NY, United States
David Choy Genentech, San Francisco, CA, United States
Sarah Pendergrass Genentech, San Francisco, CA, United States
Nizar Smaoui Abbvie, Chicago, IL, United States
Fedik Rahimov Abbvie, Chicago, IL, United States
Anne Lehtonen Abbvie, Chicago, IL, United States
Dawn Waterworth Janssen Biotech, Beerse, Belgium

**Odontology Group**
Pirkko Pussinen Hospital District of Helsinki and Uusimaa, Helsinki, Finland
Aino Salminen Hospital District of Helsinki and Uusimaa, Helsinki, Finland
Tuula Salo Hospital District of Helsinki and Uusimaa, Helsinki, Finland
David Rice Hospital District of Helsinki and Uusimaa, Helsinki, Finland
Pekka Nieminen Hospital District of Helsinki and Uusimaa, Helsinki, Finland
Ulla Palotie Hospital District of Helsinki and Uusimaa, Helsinki, Finland
Juha Sinisalo Hospital District of Helsinki and Uusimaa, Helsinki, Finland
Maria Siponen Northern Savo Hospital District, Kuopio, Finland
Liisa Suominen Northern Savo Hospital District, Kuopio, Finland

Päivi Mäntylä Northern Savo Hospital District, Kuopio, Finland
Ulvi Gursoy Hospital District of Southwest Finland, Turku, Finland
Vuokko Anttonen Northern Ostrobothnia Hospital District, Oulu, Finland
Kirsi Sipilä Northern Ostrobothnia Hospital District, Oulu, Finland
Sarah Pendergrass Genentech, San Francisco, CA, United States

**Women's Health and Reproduction Group**

Hannele Laivuori Institute for Molecular Medicine Finland, HiLIFE, University of Helsinki, Finland
Venla Kurra Pirkanmaa Hospital District, Tampere, Finland
Oskari Heikinheimo Hospital District of Helsinki and Uusimaa, Helsinki, Finland
Ilkka Kalliala Hospital District of Helsinki and Uusimaa, Helsinki, Finland
Laura Kotaniemi-Talonen Pirkanmaa Hospital District, Tampere, Finland
Kari Nieminen Pirkanmaa Hospital District, Tampere, Finland
Päivi Polo Hospital District of Southwest Finland, Turku, Finland
Kaarin Mäkikallio Hospital District of Southwest Finland, Turku, Finland
Eeva Ekholm Hospital District of Southwest Finland, Turku, Finland
Marja Vääräsmäki Northern Ostrobothnia Hospital District, Oulu, Finland
Outi Uimari Northern Ostrobothnia Hospital District, Oulu, Finland
Laure Morin-Papunen Northern Ostrobothnia Hospital District, Oulu, Finland
Marjo Tuppurainen Northern Savo Hospital District, Kuopio, Finland
Katja Kivinen Institute for Molecular Medicine Finland, HiLIFE, University of Helsinki, Finland
Elisabeth Widen Institute for Molecular Medicine Finland, HiLIFE, University of Helsinki, Finland
Taru Tukiainen Institute for Molecular Medicine Finland, HiLIFE, University of Helsinki, Finland
Mary Pat Reeve Institute for Molecular Medicine Finland, HiLIFE, University of Helsinki, Finland
Mark Daly Institute for Molecular Medicine Finland, HiLIFE, University of Helsinki, Finland
Liu Aoxing Institute for Molecular Medicine Finland, HiLIFE, University of Helsinki, Finland
Eija Laakkonen University of Jyväskylä, Jyväskylä, Finland
Niko Välimäki University of Helsinki, Helsinki, Finland
Lauri Aaltonen Hospital District of Helsinki and Uusimaa, Helsinki, Finland
Johannes Kettunen Northern Ostrobothnia Hospital District, Oulu, Finland
Mikko Arvas Finnish Red Cross Blood Service, Helsinki, Finland
Jeffrey Waring Abbvie, Chicago, IL, United States
Bridget Riley-Gillis Abbvie, Chicago, IL, United States
Mengzhen Liu Abbvie, Chicago, IL, United States
Janet Kumar GlaxoSmithKline, Brentford, United Kingdom
Kirsi Auro GlaxoSmithKline, Brentford, United Kingdom
Andrea Ganna Institute for Molecular Medicine Finland, HiLIFE, University of Helsinki, Finland
Sarah Pendergrass Genentech, San Francisco, CA, United States

**FinnGen Analysis working group**

Justin Wade Davis Abbvie, Chicago, IL, United States
Bridget Riley-Gillis Abbvie, Chicago, IL, United States
Danjuma Quarless Abbvie, Chicago, IL, United States
Fedik Rahimov Abbvie, Chicago, IL, United States
Sahar Esmaeeli Abbvie, Chicago, IL, United States
Slavé Petrovski Astra Zeneca, Cambridge, United Kingdom
Eleonor Wigmore Astra Zeneca, Cambridge, United Kingdom
Adele Mitchell Biogen, Cambridge, MA, United States
Benjamin Sun Biogen, Cambridge, MA, United States
Ellen Tsai Biogen, Cambridge, MA, United States
Denis Baird Biogen, Cambridge, MA, United States
Paola Bronson Biogen, Cambridge, MA, United States
Ruoyu Tian Biogen, Cambridge, MA, United States
Stephanie Loomis Biogen, Cambridge, MA, United States
Yunfeng Huang Biogen, Cambridge, MA, United States

Joseph Maranville Celgene, Summit, NJ, United States/ Bristol Myers Squibb, New York, NY, United States
Shameek Biswas Celgene, Summit, NJ, United States/ Bristol Myers Squibb, New York, NY, United States
Elmutaz Mohammed Celgene, Summit, NJ, United States/ Bristol Myers Squibb, New York, NY, United States
Samir Wadhawan Celgene, Summit, NJ, United States/ Bristol Myers Squibb, New York, NY, United States
Erika Kvikstad Celgene, Summit, NJ, United States/ Bristol Myers Squibb, New York, NY, United States
Minal Caliskan Celgene, Summit, NJ, United States/ Bristol Myers Squibb, New York, NY, United States
Diana Chang Genentech, San Francisco, CA, United States
Julie Hunkapiller Genentech, San Francisco, CA, United States
Tushar Bhangale Genentech, San Francisco, CA, United States
Natalie Bowers Genentech, San Francisco, CA, United States
Sarah Pendergrass Genentech, San Francisco, CA, United States
Kirill Shkura Merck, Kenilworth, NJ, United States
Victor Neduva Merck, Kenilworth, NJ, United States
Xing Chen Pfizer, New York, NY, United States
Åsa Hedman Pfizer, New York, NY, United States
Karen S King GlaxoSmithKline, Brentford, United Kingdom
Padhraig Gormley GlaxoSmithKline, Brentford, United Kingdom
Jimmy Liu GlaxoSmithKline, Brentford, United Kingdom
Clarence Wang Sanofi, Paris, France
Ethan Xu Sanofi, Paris, France
Franck Auge Sanofi, Paris, France
Clement Chatelain Sanofi, Paris, France
Deepak Rajpal Sanofi, Paris, France
Dongyu Liu Sanofi, Paris, France
Katherine Call Sanofi, Paris, France
Tai-He Xia Sanofi, Paris, France
Beryl Cummings Maze Therapeutics, San Francisco, CA, United States
Matt Brauer Maze Therapeutics, San Francisco, CA, United States
Huilei Xu Novartis, Basel, Switzerland
Amy Cole Novartis, Basel, Switzerland
Jonathan Chung Novartis, Basel, Switzerland
Jaison Jacob Novartis, Basel, Switzerland
Katrina de Lange Novartis, Basel, Switzerland
Jonas Zierer Novartis, Basel, Switzerland
Mitja Kurki Institute for Molecular Medicine Finland, HiLIFE, University of Helsinki, Finland / Broad Institute, Cambridge, MA, United States
Samuli Ripatti Institute for Molecular Medicine Finland, HiLIFE, University of Helsinki, Finland
Mark Daly Institute for Molecular Medicine Finland, HiLIFE, University of Helsinki, Finland
Juha Karjalainen Institute for Molecular Medicine Finland, HiLIFE, University of Helsinki, Finland / Broad Institute, Cambridge, MA, United States
Aki Havulinna Institute for Molecular Medicine Finland, HiLIFE, University of Helsinki, Finland
Juha Mehtonen Institute for Molecular Medicine Finland, HiLIFE, University of Helsinki, Finland
Priit Palta Institute for Molecular Medicine Finland, HiLIFE, University of Helsinki, Finland
Shabbeer Hassan Institute for Molecular Medicine Finland, HiLIFE, University of Helsinki, Finland
Pietro Della Briotta Parolo Institute for Molecular Medicine Finland, HiLIFE, University of Helsinki, Finland
Wei Zhou Broad Institute, Cambridge, MA, United States
Mutaamba Maasha Broad Institute, Cambridge, MA, United States
Shabbeer Hassan Institute for Molecular Medicine Finland, HiLIFE, University of Helsinki, Finland

Susanna Lemmelä Institute for Molecular Medicine Finland, HiLIFE, University of Helsinki, Finland
Manuel Rivas University of Stanford, Stanford, CA, United States
Aarno Palotie Institute for Molecular Medicine Finland, HiLIFE, University of Helsinki, Finland
Arto Lehisto Institute for Molecular Medicine Finland, HiLIFE, University of Helsinki, Finland
Andrea Ganna Institute for Molecular Medicine Finland, HiLIFE, University of Helsinki, Finland
Vincent Llorens Institute for Molecular Medicine Finland, HiLIFE, University of Helsinki, Finland
Hannele Laivuori Institute for Molecular Medicine Finland, HiLIFE, University of Helsinki, Finland
Sina Rüeger Institute for Molecular Medicine Finland, HiLIFE, University of Helsinki, Finland
Mari E Niemi Institute for Molecular Medicine Finland, HiLIFE, University of Helsinki, Finland
Taru Tukiainen Institute for Molecular Medicine Finland, HiLIFE, University of Helsinki, Finland
Mary Pat Reeve Institute for Molecular Medicine Finland, HiLIFE, University of Helsinki, Finland
Henrike Heyne Institute for Molecular Medicine Finland, HiLIFE, University of Helsinki, Finland
Nina Mars Institute for Molecular Medicine Finland, HiLIFE, University of Helsinki, Finland
Kimmo Palin University of Helsinki, Helsinki, Finland
Javier Garcia-Tabuenca University of Tampere, Tampere, Finland
Harri Siirtola University of Tampere, Tampere, Finland
Tuomo Kiiskinen Institute for Molecular Medicine Finland, HiLIFE, University of Helsinki, Finland

Jiwoo Lee Institute for Molecular Medicine Finland, HiLIFE, University of Helsinki, Finland / Broad Institute, Cambridge, MA, United States
Kristin Tsuo Institute for Molecular Medicine Finland, HiLIFE, University of Helsinki, Finland / Broad Institute, Cambridge, MA, United States
Amanda Elliott Institute for Molecular Medicine Finland, HiLIFE, University of Helsinki, Finland / Broad Institute, Cambridge, MA, United States
Kati Kristiansson THL Biobank / The National Institute of Health and Welfare Helsinki, Finland
Mikko Arvas Finnish Red Cross Blood Service / Finnish Hematology Registry and Clinical Biobank, Helsinki, Finland
Kati Hyvärinen Finnish Red Cross Blood Service, Helsinki, Finland
Jarmo Ritari Finnish Red Cross Blood Service, Helsinki, Finland
Miika Koskinen Helsinki Biobank / Helsinki University and Hospital District of Helsinki and Uusimaa, Helsinki
Olli Carpén Helsinki Biobank / Helsinki University and Hospital District of Helsinki and Uusimaa, Helsinki
Johannes Kettunen Northern Finland Biobank Borealis / University of Oulu / Northern Ostrobothnia Hospital District, Oulu, Finland
Katri Pylkäs University of Oulu, Oulu, Finland
Marita Kalaoja University of Oulu, Oulu, Finland
Minna Karjalainen University of Oulu, Oulu, Finland
Tuomo Mantere Northern Finland Biobank Borealis / University of Oulu / Northern Ostrobothnia Hospital District, Oulu, Finland
Eeva Kangasniemi Finnish Clinical Biobank Tampere **/** University of Tampere / Pirkanmaa Hospital District, Tampere, Finland
Sami Heikkinen University of Eastern Finland, Kuopio, Finland
Arto Mannermaa Biobank of Eastern Finland / University of Eastern Finland / Northern Savo Hospital District, Kuopio, Finland
Eija Laakkonen University of Jyväskylä, Jyväskylä, Finland
Samuel Heron University of Turku, Turku, Finland
Dhanaprakash Jambulingam University of Turku, Turku, Finland
Venkat Subramaniam Rathinakannan University of Turku, Turku, Finland
Nina Pitkänen Auria Biobank / University of Turku / Hospital District of Southwest Finland, Turku, Finland

### Biobank directors

Lila Kallio Auria Biobank / University of Turku / Hospital District of Southwest Finland, Turku, Finland
Sirpa Soini THL Biobank / The National Institute of Health and Welfare Helsinki, Finland

Jukka Partanen Finnish Red Cross Blood Service / Finnish Hematology Registry and Clinical Biobank, Helsinki, Finland
Eero Punkka Helsinki Biobank / Helsinki University and Hospital District of Helsinki and Uusimaa, Helsinki
Raisa Serpi Northern Finland Biobank Borealis / University of Oulu / Northern Ostrobothnia Hospital District, Oulu, Finland
Johanna Mäkelä Finnish Clinical Biobank Tampere / University of Tampere / Pirkanmaa Hospital District, Tampere, Finland
Veli-Matti Kosma Biobank of Eastern Finland / University of Eastern Finland / Northern Savo Hospital District, Kuopio, Finland
Teijo Kuopio Central Finland Biobank / University of Jyväskylä / Central Finland Health Care District, Jyväskylä, Finland

### FinnGen Teams

**Administration**

Anu Jalanko Institute for Molecular Medicine Finland, HiLIFE, University of Helsinki, Finland
Huei-Yi Shen Institute for Molecular Medicine Finland, HiLIFE, University of Helsinki, Finland
Risto Kajanne Institute for Molecular Medicine Finland, HiLIFE, University of Helsinki, Finland
Mervi Aavikko Institute for Molecular Medicine Finland, HiLIFE, University of Helsinki, Finland

**Analysis**

Mitja Kurki Institute for Molecular Medicine Finland, HiLIFE, University of Helsinki, Finland / Broad Institute, Cambridge, MA, United States
Juha Karjalainen Institute for Molecular Medicine Finland, HiLIFE, University of Helsinki, Finland / Broad Institute, Cambridge, MA, United States
Pietro Della Briotta Parolo Institute for Molecular Medicine Finland, HiLIFE, University of Helsinki, Finland
Sina Rüeger Institute for Molecular Medicine Finland, HiLIFE, University of Helsinki, Finland
Arto Lehisto Institute for Molecular Medicine Finland, HiLIFE, University of Helsinki, Finland
Juha Mehtonen Institute for Molecular Medicine Finland, HiLIFE, University of Helsinki, Finland
Wei Zhou Broad Institute, Cambridge, MA, United States
Masahiro Kanai Broad Institute, Cambridge, MA, United States
Mutaamba Maasha Broad Institute, Cambridge, MA, United States

**Clinical Endpoint Development**

Hannele Laivuori Institute for Molecular Medicine Finland, HiLIFE, University of Helsinki, Finland
Aki Havulinna Institute for Molecular Medicine Finland, HiLIFE, University of Helsinki, Finland
Susanna Lemmelä Institute for Molecular Medicine Finland, HiLIFE, University of Helsinki, Finland
Tuomo Kiiskinen Institute for Molecular Medicine Finland, HiLIFE, University of Helsinki, Finland
L. Elisa Lahtela Institute for Molecular Medicine Finland, HiLIFE, University of Helsinki, Finland
Matti Peura Institute for Molecular Medicine Finland, HiLIFE, University of Helsinki, Finland

**Communication**

Mari Kaunisto Institute for Molecular Medicine Finland, HiLIFE, University of Helsinki, Finland

**Data Management and IT Infrastructure**

Elina Kilpeläinen Institute for Molecular Medicine Finland, HiLIFE, University of Helsinki, Finland
Timo P. Sipilä Institute for Molecular Medicine Finland, HiLIFE, University of Helsinki, Finland
Georg Brein Institute for Molecular Medicine Finland, HiLIFE, University of Helsinki, Finland
Oluwaseun A. Dada Institute for Molecular Medicine Finland, HiLIFE, University of Helsinki, Finland
Awaisa Ghazal Institute for Molecular Medicine Finland, HiLIFE, University of Helsinki, Finland
Anastasia Shcherban Institute for Molecular Medicine Finland, HiLIFE, University of Helsinki, Finland

**Genotyping**

Kati Donner Institute for Molecular Medicine Finland, HiLIFE, University of Helsinki, Finland
Timo P. Sipilä Institute for Molecular Medicine Finland, HiLIFE, University of Helsinki, Finland

**Sample Collection Coordination**
Anu Loukola Helsinki Biobank / Helsinki University and Hospital District of Helsinki and Uusimaa, Helsinki

**Sample Logistics**
Päivi Laiho THL Biobank / The National Institute of Health and Welfare Helsinki, Finland
Tuuli Sistonen THL Biobank / The National Institute of Health and Welfare Helsinki, Finland
Essi Kaiharju THL Biobank / The National Institute of Health and Welfare Helsinki, Finland
Markku Laukkanen THL Biobank / The National Institute of Health and Welfare Helsinki, Finland
Elina Järvensivu THL Biobank / The National Institute of Health and Welfare Helsinki, Finland
Sini Lähteenmäki THL Biobank / The National Institute of Health and Welfare Helsinki, Finland
Lotta Männikkö THL Biobank / The National Institute of Health and Welfare Helsinki, Finland
Regis Wong THL Biobank / The National Institute of Health and Welfare Helsinki, Finland

**Registry Data Operations**
Hannele Mattsson THL Biobank / The National Institute of Health and Welfare Helsinki, Finland
Kati Kristiansson THL Biobank / The National Institute of Health and Welfare Helsinki, Finland
Susanna Lemmelä Institute for Molecular Medicine Finland, HiLIFE, University of Helsinki, Finland
Sami Koskelainen THL Biobank / The National Institute of Health and Welfare Helsinki, Finland
Tero Hiekkalinna THL Biobank / The National Institute of Health and Welfare Helsinki, Finland
Teemu Paajanen THL Biobank / The National Institute of Health and Welfare Helsinki, Finland

**Sequencing Informatics**
Priit Palta Institute for Molecular Medicine Finland, HiLIFE, University of Helsinki, Finland
Kalle Pärn Institute for Molecular Medicine Finland, HiLIFE, University of Helsinki, Finland
Shuang Luo Institute for Molecular Medicine Finland, HiLIFE, University of Helsinki, Finland
Shabbeer Hassan Institute for Molecular Medicine Finland, HiLIFE, University of Helsinki, Finland

**Trajectory Team**
Tarja Laitinen Pirkanmaa Hospital District, Tampere, Finland
Harri Siirtola University of Tampere, Tampere, Finland
Javier Gracia-Tabuenca University of Tampere, Tampere, Finland

**Data protection officer**
Tero Jyrhämä Institute for Molecular Medicine Finland, HiLIFE, University of Helsinki, Finland

**FinBB - Finnish biobank cooperative**
Marco Hautalahti
Laura Mustaniemi
Mirkka Koivusalo
Sarah Smith
Tom Southerington
